# Past Disparities in Advance Care Planning Across Sociodemographic Characteristics and Cognition Levels in the United States

**DOI:** 10.1101/2024.05.09.24307125

**Authors:** Zahra Rahemi, Juanita-Dawne R. Bacsu, Sophia Z. Shalhout, Morteza Sabet, Delaram Sirizi, Matthew Lee Smith, Swann Arp Adams

## Abstract

We aimed to examine past advance care planning (ACP) in U.S. older adults across different sociodemographic characteristics and cognition levels. We established the baseline trends from 10 years ago to assess if trends in 2024 have improved upon future data availability. We considered two legal documents in the Health and Retirement Study 2014 survey as measures for ACP: a living will and durable power of attorney for healthcare (DPOAH). Logistic regression models were fitted with outcome variables (living will, DPOAH, and both) stratified by cognition levels (dementia/impaired cognition versus normal cognition). Predictor variables included age, gender, ethnicity, race, education, marital status, rurality, everyday discrimination, social support, and loneliness. Age, ethnicity, race, education, and rurality were significant predictors of ACP (having a living will, DPOAH, and both the living will and DPOAH) across cognition levels. Participants who were younger, Hispanic, Black, had lower levels of education, or resided in rural areas were less likely to complete ACP. Examining ACP and its linkages to specific social determinants is essential to understanding disparities and educational strategies needed to facilitate ACP uptake among different population groups. Accordingly, this study aimed to examine past ACP disparities in relation to specific social determinants of health and different cognition levels. Future studies are required to evaluate whether existing disparities have improved over the last 10 years when 2024 data is released. Addressing ACP disparities among diverse populations, including racial and ethnic minorities with reduced cognition levels, is crucial for enhancing health equity and access to care.

## Introduction

Among adults aged 65 and older in the United States (U.S.), Alzheimer’s disease and related dementias (ADRD) are the fifth leading cause of death. The prevalence of ADRD in the U.S. is projected to reach approximately 14 million by the year 2050 [1]. The prevalence and challenges of ADRD differ by ethnicity [2], rurality [3, 4] gender, education, age, and co-morbidities [5]. For instance, this prevalence is elevated among individuals of Hispanic ethnicity, residing in rural areas, identifying as female, having lower levels of education, being of older age, and having co-morbidities. Overall, older adults from underrepresented and racial/ethnic minority groups have higher rates of chronic disease and cognitive impairment and lower rates of advance care planning (ACP) and comfort care at the end of life (EOL) [3, 6, 7]. It is imperative to explore the underlying causes of disparities in ACP and EOL care, aiming to eradicate inequities in healthcare based on factors such as race, ethnicity, and other personal identities.

In the context of ADRD patients, ACP and EOL care present distinct challenges [7]. As dementia progresses and decisional capacity diminishes, healthcare decisions often fall to a family member or proxy. This can be particularly challenging when life expectancy remains uncertain and ACP discussions have not taken place [8]. ACP is a process across different levels of health and encompasses both formal mechanisms or written documents—such as advance directives, living will, and durable power of attorney for healthcare (DPOAH)—and informal mechanisms— including verbal discussions of healthcare preferences with family and providers [9, 10]. ACP facilitates proactive communication of healthcare decisions before impairment of decision-making capacity. Evidence indicates ACP improves the quality of care, communication satisfaction, and patient care satisfaction among patients, surrogates, and healthcare providers. ACP also reduces distress for surrogates and healthcare providers while promoting decision-making aligned with patients’ values and goals [9, 11]. ACP holds particular significance for individuals experiencing cognitive impairment, as their gradual loss of decision-making capacity, along with issues related to EOL, may present challenges for both their family members and proxy decision-makers.

This study used the 2014 Health and Retirement Study (HRS) data to examine past ACP in U.S. older adults across different sociodemographic characteristics and cognition levels. Compared to its preceding waves, we chose the 2014 wave because it offered the most comprehensive data among the selected harmonized datasets, particularly in End-of-Life (EOL) sections. Our overarching objective is to replicate the study using the HRS 2024 data upon its release, enabling us to assess ACP disparities over a span of 10 years. We are particularly interested in understanding how ACP disparities may change over time. Utilizing HRS waves aids in the completion of promising ongoing studies and leverages long-term data to provide valuable insights for establishing best practices in addressing health disparities, including topics related to EOL disparities.

Existing studies document trends of disparities over time among marginalized groups in relation to ACP. More specifically, structural disparities in accessing ACP have included issues of institutional racism, implicit bias, practitioner discomfort in discussing EOL planning, language differences, lack of cultural awareness (different beliefs and values) [12], exacerbation of institutional barriers/constraints, and pre-conceived notions of patients’ EOL wishes [13]. Moreover, healthcare providers are documented as avoiding ACP conversations with patients from racial and/or ethnic minority groups (such as Black, Asian, Hispanic, and Native American), non-English speaking patients with low incomes, and individuals with low health literacy [12]. Individual-level issues related to disparities included lack of ACP awareness and knowledge; inability to make decisions; no surrogates available; patient discomfort in discussing ACP (religious, cultural, or individual factors); lack of trust in healthcare providers and the healthcare system [14]; family involvement; financial challenges; and faith and religious beliefs [13]. Despite this knowledge, existing studies do not examine past ACP disparities in relation to different cognition levels. However, evaluating changes in ACP disparities over time among marginalized population groups, including different cognition levels is crucial for improving health equity and access to care, especially for people living with dementia.

We defined ACP as encompassing a living will and DPOAH. A living will is a document that articulates a person’s preferences for medical treatments, offering insight into the healthcare they wish to receive (or decline) during severe health conditions or when their medical decision-making capacity is impaired. A DPOAH designates a person to make healthcare decisions on behalf of another individual when they are incapable of making decisions themselves [10]. The theoretical framework underpinning this research is the end-of-life care planning model, a nursing conceptual model that provides a framework for understanding factors associated with ACP disparities in diverse individuals [15].

## Materials and Methods

### Design and Setting

We conducted an observational, cross-sectional study utilizing the HRS, a comprehensive nationally representative dataset that encompasses information from over 43,000 respondents. The HRS team employs a probability sample method, with oversampling of African American, Hispanic, and Floridian participants, to gather information from adults aged 51 years and older every two years. This data collection covers various aspects, including health, functional status, family structure, demographics, and lifestyle activities [16].

Our analyses used a sample of 17,698 participants from the HRS 2014 survey (Wave 12). To ensure adequate statistical power and examine relationships between study variables, we combined data from two different HRS datasets to create a pooled cross-sectional dataset: the harmonized HRS version B and the 1992-2016 Rand HRS Longitudinal version 2. Sub-data files were generated from the original files, and subsequent procedures, such as data cleaning, computing, and appropriate variable recoding, were performed. The final dataset was then created by merging all subfiles.

### Variables

Race was captured as white, black, or other. Ethnicity was designated as Hispanic versus non-Hispanic. Education was categorized as less than or equal to a high school education and greater than a high school education. Rurality was grouped as an urban or rural residence. Marital status was designated as married/partnered or not married/divorced/widowed/single.

### Data Analysis

Data analyses were performed using SAS Version 9.4. Descriptive statistics were computed to explore the frequencies and distributions of key variables. Given the HRS’s complex sampling design, which includes oversampling of minorities and multiple respondents from a household, individual and household weights were utilized for some analyses. Descriptive statistics were analyzed using weighted methods (Proc SurveyMeans, Proc SurveyFreq) to account for survey design complexities. However, to avoid potential bias in the inferential model analyses [17], we employed unweighted regression modeling procedures (Proc Logistic).

The primary outcomes modeled were whether the respondents had a living will (“whether respondent has a living will” [1= yes, 0= no]) and a DPOAH (“whether respondent has durable power of attorney” [1= yes, 0= no]). A summary joint outcome variable was created from these two variables (having both ACP measures, at least one, and none). Predictor variables included age at interview, gender, ethnicity, race, education, marital status, rurality, everyday discrimination score, social support score (both spousal and friends), and loneliness score.

We adopted the Langa-Weir approach, a composite measure that ranges from 0 to 27, to distinguish cognitive impairment from more advanced dementia [18]. In this study, the cohort was split into two groups: dementia/impaired cognition (Langa-Weir score < 11) and normal cognition (Langa-Weir score > 11). Logistic regression models were run with each outcome variable (having living will DPOAH, and both the living will and DPOAH) stratified by cognition (dementia/impaired cognition versus normal cognition). Since the combination variable (both living will and DPOAH) had 3 levels, we utilized polytomous logistic regression models. In the full model for each outcome, we incorporated all predictor variables that were significant in the univariate models.

## Results

Of the 17,698 respondents, 77.8% had normal cognition, and 22.2% had a diagnosis of dementia/impaired cognition. Participants who were older, Hispanic, Black, single/widowed (no partner), had less education, and had higher loneliness scores were more likely to have dementia/impaired cognition than their counterparts (Table 1). Those with impaired cognition or dementia had significantly higher spousal support compared to those without dementia (p<0.01) but significantly lower other family social support (p<0.01). Additionally, among those with dementia or impaired cognition, only 46.2% had a living compared to 54.2% among normal cognition individuals (p<0.01). Similarly, only 50.6% of those individuals with dementia or impaired cognition had a durable power of attorney versus 54.2% of those individuals with normal cognition (p<0.03). Among those with dementia or impaired cognition, 38.7% had both a DPOA and living will compared to 47.5% among normal cognition individuals (p<0.01).

**Table 1.** Descriptive Statistics for the 2014 Health and Retirement Survey Cohort by Cognition Group (HRS, 2014)

|  | Normal Cognition<br>N=13,774<br>Weighted % (n) | Dementia/Impaired Cognition<br>N=3924<br>Weighted % (n) | P-Value |
| --- | --- | --- | --- |
| Education |  |  |  |
| ≤ 12 years | 38.8 (5572) | 72.8 (2733) | < 0.01 |
| 13+ years | 61.2 (7062) | 27.2 (890) |  |
| Gender |  |  |  |
| Male | 45.1 (5304) | 45.4 (1517) | 0.05 |
| Female | 54.9 (7494) | 54.6 (2124) |  |
| Race |  |  |  |
| White | 86.3 (9666) | 66.9 (2084) | < 0.01 |
| Black/Other | 13.7 (2999) | 33.1 (1546) |  |
| Ethnicity |  |  |  |
| Hispanic | 7.1 (1444) | 17.0 (752) | < 0.01 |
| Non-Hispanic | 92.9 (11237) | 83.0 (2882) |  |
| Marital Status |  |  |  |
| Married/Living with a Partner | 76.0 (9743) | 70.4 (2708) | <0.01 |
| Single/Widowed | 24.0 (2952) | 29.6 (931) |  |
| Rurality |  |  |  |
| Rural | 26.4 (3163) | 28.3 (937) | 0.27 |
| Urban | 73.6 (9480) | 71.7 (2678) |  |
| Living Will |  |  |  |
| Yes | 54.2 (3743) | 46.2 (1071) | <0.01 |
| No | 45.8 (3051) | 53.8 (1396) |  |
| Durable Power of Attorney |  |  |  |
| Yes | 54.2 (3787) | 50.6 (1215) | <0.03 |
| No | 45.8 (3024) | 49.4 (1252) |  |
| Joint Effects |  |  |  |
| Both DPOA & Living Will | 47.5 (3259) | 38.7 (892) | <0.01 |
| Either DPOA or Living Will | 13.3 (956) | 19.1 (470) |  |
| None | 39.2 (2547) | 42.2 (1078) |  |
|  | **Mean (SE)** | **Mean (SE)** |  |
| Age (yrs) | 65.7 (0.3) | 72.3 (0.4) | < 0.01 |
| Everyday Discrimination Score | 1.54 (0.01) | 1.59 (0.02) | 0.07 |
| Spousal Social Support Score | 1.72 (0.01) | 1.86 (0.03) | < 0.01 |
| Other Family Social Support Score | 1.82 (0.01) | 1.76 (0.02) | < 0.01 |
| Loneliness Score | 1.50 (0.01) | 1.63 (0.02) | < 0.01 |

The significant predictors of having a living, stratified by cognition status, are shown in Table 2. For those individuals with dementia or impaired cognition, race, ethnicity, education, and age significantly predicated having a living will such that black persons were 0.36 (95% CI: 0.21, 0.61) times less likely, Hispanic persons were 0.34 (95% CI: 0.19, 0.63) times less likely, and those with more than a high school education were 2.27 (95% CI: 1.43, 3.62) times more likely. The odds of having a living will increased by 7% for each 1 year increase in age among persons with dementia or impaired cognition. Among those with normal cognition, race (OR_black_=0.43, 95% CI: 0.30-0.63), ethnicity (OR_Hispanic_=0.30, 95% CI: 0.19-0.47), rurality (OR_rural_=0.81, 95% CI: 0.65, 0.99), education (OR_>HS_= 1.85, 95% CI: 1.51-2.25), and age (OR=1.07, 95% CI: 1.05-1.09) were significant predictors of having a living will.

**Table 2.** Living Will Prediction Model.

|  |  | Dementia/Impaired Cognition<br>N=517 |  | Normal Cognition<br>N=1850 |  |
| --- | --- | --- | --- | --- | --- |
|  |  | Odds Ratio | 95% CI | Odds Ratio | 95% CI |
| Race | White | 1.00 |  | 1.00 |  |
|  | Black | **0.36** | **0.21, 0.61** | **0.43** | **0.30, 0.63** |
|  | Other | 0.78 | 0.33, 1.83 | 0.87 | 0.50, 1.50 |
| Ethnicity | Not Hispanic | 1.00 |  | 1.00 |  |
|  | Hispanic | **0.34** | **0.19, 0.63** | **0.30** | **0.19, 0.47** |
| Rural | No | 1.00 |  | 1.00 |  |
|  | Yes | 0.83 | 0.54, 1.27 | **0.81** | **0.65, 0.99** |
| Marital Status | Single | 1.00 |  | 1.00 |  |
|  | Married | 1.36 | 0.58, 3.21 | 0.88 | 0.55, 1.40 |
| Gender | Male | 1.00 |  | 1.00 |  |
|  | Female | 0.98 | 0.66, 1.44 | 1.13 | 0.93, 1.39 |
| Education | ≤ HS | 1.00 |  | 1.00 |  |
|  | >HS | **2.27** | **1.43, 3.62** | **1.85** | **1.51, 2.25** |
| Age |  | **1.07** | **1.04, 1.11** | **1.07** | **1.05, 1.09** |
| Everyday Discrimination Score |  | 1.14 | 0.86, 1.51 | 1.05 | 0.88, 1.26 |
| Other Support |  | 0.82 | 0.56, 1.20 | 1.02 | 0.84, 1.25 |
| Spousal Support |  | 1.01 | 0.68, 1.51 | 0.94 | 0.77, 1.15 |
| Loneliness Score |  | 1.16 | 0.69, 1.94 | 0.78 | 0.60, 1.02 |

Note. Results in bold indicate significance at a significance level of 0.05.

Similar findings were demonstrated for DPOA (Table 3). Among individuals with dementia or impaired cognition, only education (OR_>HS_=1.96, 95% CI: 1.25-3.06) and age (OR=1.05, 95% CI: 1.02-1.08) were significant predictors of DPOA. Among those with normal cognition, race (OR_black_=0.64, 95% CI: 0.44-0.92), ethnicity (OR_Hispanic_=0.43, 95% CI: 0.28-0.66), education (OR_>HS_= 1.85, 95% CI: 1.52-2.26), and age (OR=1.07, 95% CI: 1.05-1.08) were significant predictors.

**Table 3.** Durable Power of Attorney Prediction Model.

|  |  | Dementia/Impaired Cognition<br>N=517 |  | Normal Cognition<br>N=1844 |  |
| --- | --- | --- | --- | --- | --- |
|  |  | Odds Ratio | 95% CI | Odds Ratio | 95% CI |
| Race | White | 1.00 |  | 1.00 |  |
|  | Black | 0.69 | 0.43, 1.11 | **0.64** | **0.44, 0.92** |
|  | Other | 1.56 | 0.72, 3.35 | 0.79 | 0.46, 1.35 |
| Ethnicity | Not Hispanic | 1.00 |  | 1.00 |  |
|  | Hispanic | 0.60 | 0.34, 1.04 | **0.43** | **0.28, 0.66** |
| Rural | No | 1.00 |  | 1.00 |  |
|  | Yes | 1.16 | 0.78, 1.73 | 0.82 | 0.66, 1.01 |
| Marital Status | Single | 1.00 |  | 1.00 |  |
|  | Married | 1.59 | 0.73, 3.46 | 0.95 | 0.60, 1.50 |
| Gender | Male | 1.00 |  | 1.00 |  |
|  | Female | 1.02 | 0.70, 1.48 | 1.17 | 0.96, 1.43 |
| Education | ≤ HS | 1.00 |  | 1.00 |  |
|  | >HS | **1.96** | **1.25, 3.06** | **1.85** | **1.52, 2.26** |
| Age |  | **1.05** | **1.02, 1.08** | **1.07** | **1.05, 1.08** |
| Everyday Discrimination Score |  | 0.93 | 0.72, 1.21 | 1.07 | 0.89, 1.27 |
| Other Support |  | 1.02 | 0.71, 1.45 | 1.05 | 0.86, 1.27 |
| Spousal Support |  | 0.98 | 0.67, 1.42 | 0.83 | 0.68, 1.01 |
| Loneliness Score |  | 1.30 | 0.80, 2.09 | 0.87 | 0.66, 1.13 |

Note. Results in bold indicate significance at a significance level of 0.05.

**Table 4.** Joint Effects of Living Will and Durable Power of Attorney Prediction Model.

|  |  | Dementia/Impaired Cognition<br>N=511 |  | Normal Cognition<br>N=1844 |  |
| --- | --- | --- | --- | --- | --- |
|  |  | OR (95% CI)<br>Both LW/DPOA | OR (95% CI)<br>Just One ACM* | Odds Ratio<br>Both LW/DPOA | 95% CI<br>Just One ACM* |
| Race | White | 1.00 | 1.00 | 1.00 | 1.00 |
|  | Black | **0.38 (0.21, 0.69)** | 1.36 (0.74, 2.52) | **0.46 (0.30, 0.70)** | 1.19 (0.73, 1.94) |
|  | Other | 1.05 (0.40, 2.73) | 1.71 (0.66, 4.44) | 0.81 (0.45, 1.45) | 0.77 (0.32, 1.84) |
| Ethnicity | Not Hispanic | 1.00 | 1.00 | 1.00 | 1.00 |
|  | Hispanic | **0.36 (0.18, 0.70)** | 0.82 (0.39, 1.70) | **0.32 (0.20, 0.52)** | **0.40 (0.20, 0.82)** |
| Rural | No | 1.00 | 1.00 | 1.00 | 1.00 |
|  | Yes | 0.93 (0.58, 1.48) | 1.36 (0.80, 2.32) | **0.79 (0.63, 0.99)** | 1.12 (0.82, 1.53) |
| Marital Status | Single | 1.00 | 1.00 | 1.00 | 1.00 |
|  | Married | 1.73 (0.63, 4.75) | 1.11 (0.44, 2.75) | 0.90 (0.55, 1.48) | 0.99 (0.49, 2.00) |
| Gender | Male | 1.00 | 1.00 | 1.00 | 1.00 |
|  | Female | 1.02 (0.66, 1.57) | 0.79 (0.48, 1.30) | 1.19 (0.96, 1.48) | 1.18 (0.87, 1.60) |
| Education | ≤ HS | 1.00 | 1.00 | 1.00 | 1.00 |
|  | >HS | **2.65 (1.56, 4.51)** | **2.61 (1.43, 4.79)** | **2.07 (1.67, 2.57)** | **1.37 (1.02, 1.85)** |
| Age |  | **1.08 (1.04, 1.12)** | **1.05 (1.01, 1.10)** | **1.08 (1.06, 1.10)** | **1.07 (1.04, 1.10)** |
| Everyday Discrimination Score |  | 1.01 (0.73, 1.39) | 1.29 (0.93, 1.78) | 1.07 (0.88, 1.31) | 1.17 (0.90, 1.51) |
| Other Support |  | 0.87 (0.56, 1.33) | 1.42 (0.90, 2.25) | 1.04 (0.84, 1.30) | 1.33 (0.99, 1.79) |
| Spousal Support |  | 1.05 (0.68, 1.63) | 0.60 (0.35, 1.03) | 0.85 (0.68, 1.06) | 0.89 (0.65, 1.21) |
| Loneliness Score |  | 1.22 (0.69, 2.17) | 1.43 (0.76, 2.68) | 0.79 (0.59, 1.06) | 0.69 (0.45, 1.05) |

Note. \* ACM- Advanced Care Measure (either LW or DPOA). Results in bold indicate significance at a significance level of 0.05.

In models examining the joint effects of having a living will or DPOA, higher education and age significantly predicted an increased odds of having both or at least one ACP for both impaired cognition/dementia individuals and normal cognition individuals. Black race and Hispanic ethnicity predicted a lower likelihood of having both ACP but not just one ACP for both cognition groups. Living in a rural area was only a significant predictor of the decreased likelihood of having both ACP measures among normal cognition individuals.

## Discussion

We examined the involvement of older adults in ACP (having a living will and DPOAH) and identified the factors associated with ACP among participants with different cognitive statuses. Consistent with the end-of-life care planning model [15], our findings revealed ACP disparities based on sociodemographic factors and cognition levels. Individuals living with cognitive impairments and with specific identities, such as Hispanic and Black participants, those with lower education levels, and residents of rural areas, had lower rates of ACP. Our results showed ACP disparities among diverse populations.

Cognitive impairments and ADRD are progressive conditions, and patients gradually lose their decision-making ability and cognitive function. Therefore, early healthcare decision-making and planning through ACP can significantly enhance the quality of care for individuals with cognitive impairments. Moreover, ACP’s proactive approach reduces anxiety about last-minute decisions for patients and their caregivers [9, 11]. There are multiple barriers to initiating ACP in individuals with ADRD including topic avoidance, stigma/fear of diagnosis, denial and family disagreements, inadequate knowledge, ADRD behavioral changes, dementia trajectory, and nihilistic views that nothing can be done [7, 8, 19–21]. Additionally, research suggests that ACP barriers encompass various sociodemographic factors, living and residential conditions, acculturation, spirituality, religiosity, absence of a proxy for appointment if required, and cultural beliefs [22–25]. Consistent with our results, racial/ethnic minority, culturally diverse, and rural populations experience disparities in ACP and EOL care [4, 23, 24, 26]. Engagement in ACP is less common among African Americans and Hispanic Americans, and they are more inclined to prefer intensive and curative treatments over comfort care at EOL [24, 25].

Research indicates that the lack of ACP can lead to reduced utilization of supportive care, higher rates of curative interventions, increased stress for caregivers, and diminished care satisfaction [8, 9, 11]. ACP is crucial to enhancing shared decision-making and facilitating discussions about EOL care options, including hospice services. It enables care management that aligns with the patient’s and their family’s preferences, contributing to improved patient dignity [4, 27]. Improving ACP engagement is especially important for people with cognitive impairment before they lose functional, cognitive, and decisional capacity. This study focused on understanding ACP disparities among diverse participants across different cognition levels and identities in the past (i.e., 10 years ago) to serve as a baseline for comparison today upon release of the 2024 data. This will allow us to determine improvements, if any, in trends. Our findings may further EOL-related research, practice, and policy by informing the development of dementia-specific and person-centered ACP methods. The results can incorporate the body of knowledge to be used for reducing racial and ethnic disparities related to ACP and EOL care.

## Limitations

Our study was limited to secondary data analyses, as it drew from existing data and measurement tools. However, a strength of the study was its reliance on a nationally representative survey of U.S. adults, which minimized the potential for sampling bias and threats to external validity.

Additionally, the HRS, with an oversampling of Black and Hispanic older adults, helps investigate and address racial and ethnic health disparities across the U.S.

## Conclusion

Our study identified and quantified ACP disparities among older adults with cognitive impairment and diverse identities, especially in terms of ethnicity/race, rurality, and decreased educational attainment. Our findings provide valuable information for shaping educational interventions to support EOL planning for diverse older adults. Moreover, understanding ACP barriers can aid clinicians in having proactive ACP discussions with their patients living with ADRD and their caregivers. The findings can also help understand and eliminate ACP and EOL care disparities based on race and ethnicity. Overall, our findings can inform research, practice, and policy that works to enhance ACP in diverse populations with cognitive impairment. The knowledge of ACP disparities and their related factors is pivotal for healthcare professionals to design methods to reduce disparities in ACP and EOL care. It will be of interest to the research community to understand whether disparities have improved over a decade following the release of the 2024 data. Replicating this study could offer valuable insights. Continued studies utilizing long-term datasets will contribute to our understanding of the underlying factors that contribute to perpetuating healthcare and EOL disparities.

## Data Availability

All data produced in the present study are available upon reasonable request to the authors

## Statements and Declarations

### Author Contributions

All authors contributed to the study’s conception and design. Data preparation was performed by Zahra Rahemi and Swann Adams. Data analysis was collected by Swann Adams. The results were interpreted by Zahra Rahemi and Juanita-Dawne Bacsu. The first draft of the manuscript was written by Zahra Rahemi, Morteza Sabet, Delaram Sirizi, and Matthew Lee Smith. Sophia Shalhout, the first author’s mentor, supervised all the steps and participated in finalizing the manuscript. All authors commented on previous versions of the manuscript. All authors read and approved the final manuscript.

### Ethics Approval

Clemson University Institutional Review Board exempted the study protocols (IRB Number: IRB2021-0720).

